# Global trend and disparity in the burden of ischemic stroke attributable to high body-mass index from 1990 to 2021 and projection to 2050: a systematic analysis based on the Global Burden of Disease Study 2021

**DOI:** 10.1101/2025.11.26.25341126

**Authors:** Shuting Ni, Ying Zhang, Yongxiang Zhang, Shilei Lin, Tong Zhao

**Author notes:** Corresponding Author: *Tong Zhao. These authors contributed equally to this work (Shuting Ni, Ying Zhang, Yongxiang Zhang).

## Abstract

**Background:** Obesity and overweight are increasingly recognized as significant risk factors for the incidence and progression of ischemic stroke (IS). However, its epidemiological investigation including the disease burden and its trends remains insufficiently explored. This research aimed to reveal and predict the disease burden of ischemic stroke attributable to high body-mass index (IS-HBMI), which would offer significant references for focused prevention and disease management methods.

**Methods:** The study extracted data from the Global Burden of Disease Study 2021 (GBD 2021). Death cases, disability-adjusted life years (DALYs) case, age-standardized mortality rate (ASMR) and age-standardized DALYs rate (ASDR) were obtained from GBD 2021 to assess the global burden from 1990 to 2021. Decomposition analysis explored the driving factors to IS-HBMI. The Average Annual Percent Change (AAPC) in ASMR and ASDR of IS-HBMI was determined to analyze temporal trends by Joinpoint regression analysis. Bayesian age-period-cohort (BAPC) modelling was used to predict the burden of disease up to 2050.

**Results:** The global deaths and DALYs of IS-HBMI were 4439186 and 51520000 in 2021, exhibiting a continuous growth trend over the past 32 years. The ASMR and ASDR for males showed faster growth. The disease burden was greatest among middle-aged and older populations, while the rapidly increase in adolescents should not be overlooked. High sociodemographic index (SDI) regions and Latin America each recorded the highest disease burden within their respective categories of SDI regions and GBD regions. Additionally, Predictive models indicated a gradual upward trend from 2022 to 2049.

**Conclusion:** The study revealed that the global disease burden of IS-HBMI had continuously increased from 1990 to 2021, and it was predicted to escalate until 2050. The findings emphasize the need for more detailed IS screening and weight loss measures tailored to specific regions and populations, which would benefit efforts to curb the projected rise in IS-HBMI deaths.

## Introduction

Ischemic stroke is a neurological dysfunction caused by a vascular accident and is an important contributor to disability and death[1]. According to the latest data, there are approximately 7.8 million new cases of IS worldwide each year, accounting for 62.4 % of all stroke cases worldwide [2, 3]. IS triggers a complex series of ischemic cascade reactions, ultimately resulting in irreversible neuronal damage and cerebral infarction. This process can result in devastating dysfunction for the individual patient, as well as a heavy financial and caregiving burden on his or her family and society. The rapid progression of ischemic stroke can be attributed to a number of factors, the main risk factor being hypertension [4]. In addition, many studies have highlighted risk factors such as high cholesterol, diabetes, obesity, and end-stage renal disease[5–9]. Many of these risk factors are strongly associated with weight gain[10].

In recent years, more and more studies have focused on the relationship between obesity/overweight and IS, emphasising that obesity/overweight is an important risk factor for the development of IS. A study suggests that stroke risk rises significantly when BMI >25 kg/m² (95% CI: 1.45 to 1.79)[11]. According to the results of this study, the average annual percentage change (AAPC) in age-standardized mortality rate (ASR, referring to age-standardized DALYs rate) of IS-HBMI worldwide from 1990 to 2021 was −0.58, with −0.1 in males and −0.92 in females[10].A prospective cohort study also found a significant association between high BMI and perioperative ischemic stroke[12].Other studies, including Mendelian Randomization analyses and meta-analyses, have further reinforced the significant correlation between obesity/overweight and the development of IS[13]. In addition, several studies have further demonstrated that factors such as hyperinsulinaemia, chronic inflammation, disorders of lipid metabolism and hormonal changes contribute to the increased susceptibility to IS in obese/overweight individuals[14, 15].

Although previous studies have revealed an association between obesity/overweight and IS, there is still a lack of comprehensive study on the burden of ischemic stroke attributable to high body-mass index (IS-HBMI) and its trends across sexes, age groups, regions, and countries/territories. This study comprehensively utilized data from the Global Burden of Disease Study 2021 (GBD 2021) to analyze the trends and disparities in the burden of IS-HBMI from 1990 to 2021 and predicted its future trajectory up to 2050, which was expected to provide a scientific foundation for future targeted prevention and intervention strategies.

## Methods

The GBD 2021, led by the Institute for Health Metrics and Evaluation (IHME) at the University of Washington, employs standardized and comparable methods to assess disease burden data across 204 countries and regions, covering 371 diseases and injuries and 88 risk factors [16, 17]. All data are publicly available at https://vizhub.healthdata.org/gbd-results/. The GBD 2021 originates from various credible official sources, encompassing regional and national censuses, vital statistics, disease registries, and other authoritative databases. Data identification, extraction, and integration are rigorously performed through systematic assessments of published studies, alongside information from websites, reports and primary datasets from governments, international organizations and collaborators. Stringent quality control, evaluation, and calculation procedures are implemented to guarantee data accuracy. This study used publicly accessible data from GBD 2021, which has been approved by the ethical committee of the Institute for Health Metrics and Evaluation (IHME). Thus, no additional ethical approval was required, and the study adhered to the Guidelines for Accurate and Transparent Health Estimates Reporting (GATHER) for cross-sectional studies[18].

For IS, the International Classification of Diseases (ICD-10) codes are I63.x, G46.3, and G46.4[19]. BMI is a measure of body weight, calculated by an individual’s dividing weight (in kilograms) by the square of height (in meters)[20].It is commonly used for the primary diagnosis of obesity and overweight due to its simple calculation method. And it is also widely employed to assess nutritional status, body composition and physical development levels. According to the BMI classification, weight categories are defined as follows: underweight (BMI below 18.5), normal weight range (18.5 to 24.9), overweight (25.0 to 29.9), and obesity (greater than30.0). In this study, ‘High BMI’ refers to a BMI of 25 or above[21].

The burden of IS-HBMI was assessed using absolute numbers of deaths and disability-adjusted life years (DALYs), as well as their corresponding age-standardized rates. Specifically, we utilized two key metrics:The age-standardized mortality rate (ASMR). The age-standardized DALY rate (ASDR). DALYs, a summary measure of overall disease burden, combine the years of life lost due to premature mortality (YLLs) and the years lived with disability (YLDs). Both absolute numbers and age-standardized rates (ASMR and ASDR) for IS-HBMI were obtained directly from the GBD 2021 study. All estimates are presented with their 95% uncertainty intervals (UIs).

In examining disease burden in various countries/territories, we also employed the socio-demographic Index (SDI) to assess the level of socioeconomic development of a country or region. GBD 2021 divides countries/territories into five stages of socioeconomic development: high (SDI > 0.81), middle-high (SDI between 0.70 to 0.81), middle (SDI between 0.61 to 0.69), middle-low (SDI between 0.46 to 0.60), and low (SDI < 0.46). Based on this, differences in disease burden of IS-HBMI among countries/territories with differing degrees of economic development could be investigated, allowing for the establishment of more precise preventative measures adapted to the economic development context of each country/territory[22].

Joinpoint regression analysis was used to conduct in-depth analysis of disease burden trends. The software of Joinpoint was developed by the National Cancer Institute’s Division of Cancer Control and Population Sciences, which could be downloaded from the following website: https://surveillance.cancer.gov/joinpoint/. It can identify significant time points of change in global ASMR and ASDR from 1990 to 2021. The average annual percent change (AAPC) describes the average annual percent change in ASR over 32 years, whereas the annual percent change (APC) quantifies the percent change in ASR for a certain time period [23].

A prediction study was developed using Bayesian age-period-cohort (BAPC) model, in which Bayesian statistical techniques were applied to analyze correlations between age, period, and cohort in demographic data[24]. This method has been widely applied in demographic and epidemiological study, and it has previously been used to predict disease burden patterns using historical data[25]. BAPC model utilizes integrated nested Laplace approximations (INLA) for complete Bayesian inference, calculating both age-specific and age-standardized predicted rates, while automatically incorporating Poisson noise when focusing on the predicted distribution. Previous studies have shown that the BAPC model has a relatively lower error rate compared to other prediction models, which can more accurately and comprehensively predict the trends in disease burden changes[26]. All statistical analysis and mapping were conducted by R software (version 4.3.2). All statistical tests were two sided, and P-values less than 0.05 were considered statistically significant.

## Results

### 1. Global disease burden of IS-HBMI in 2021

In 2021, the global number of ischemic stroke (IS-HBMI) deaths attributable to high body mass index (HBMI) was 4439186 (95% UIs: 64903-8647485), an increase of 208.8% compared with 1990, whereas the age-standardised mortality rate (ASR) showed a significant trend of improvement, from 55.3/100,000 population [95% UIs:7.9-109.16] to 51.52/100,000 population [95% UIs: 7.52-100.28]. Joinpoint regression analysis showed an average annual percentage change (AAPC) in ASR of −0.58, with the fastest rate of decline from 2001-2010 (APC=-2.01), and significant regional heterogeneity: the AAPC of −1.49 in high SDI areas was significantly different from that in low SDI areas (−1.47) (p<0.001). Although the absolute difference in AAPC between high and low SDI regions was small, the statistical significance (p < 0.001) indicates a robust difference in the trends, potentially driven by large sample sizes and narrow confidence intervals. Decomposition analyses further revealed a triple divergence in the driving mechanisms: the effect of population growth was the primary factor in the increase in deaths (61.2% of the total contribution, with 72.5% in the low SDI regions), and the effect of ageing contributed negatively (−18.7% of the total), with high SDI regions (e.g., high-income Asia and the Pacific, Western Europe) supporting the global decline in the risk of deaths significantly more than low SDI regions (only −0.47 (95% CI: −0.52 to −0.43)) and 3% more than low SDI regions (−11.8%)(fig 4). High SDI regions supported the global DALYs risk reduction with AAPC=-1.49, which was consistent with that of low SDI regions (AAPC=-1.47) (p<0.001), while the epidemiological transition effect (−20.3% of the total share) played a central mitigating role. These findings highlight a divergence in the global burden of IS-HBMI in terms of absolute numbers and declining standardisation rates. Specifically, population growth in low-SDI regions contributed significantly more to mortality (72.5%) than the global average (61.2%), serving as the primary driver of increased burden; Meanwhile, the negative contribution of aging in high SDI regions (−18.7%) is more pronounced, and its supportive role in reducing global mortality risk (AAPC=-3.74) is significantly stronger than that in low SDI regions (AAPC=-1.47). This disparity clearly reflects the heterogeneous driving mechanisms of IS-HBMI burden across different developmental stages, indicating substantial socioeconomic differences in IS-HBMI burden The global burden of disease for IS-HBMI in 2021 showed significant gender differences: 2111409 deaths [95% UIs, 307233-4133989, ASR=51.98 [7.53-101.35]] in men and 2327777 deaths [95% UIs, 3416.68-45069.72, ASR=50.74 [7.46-98.07]] in women, with male AS R significantly higher than female (p<0.001) (fig.1).

**Fig 1.**
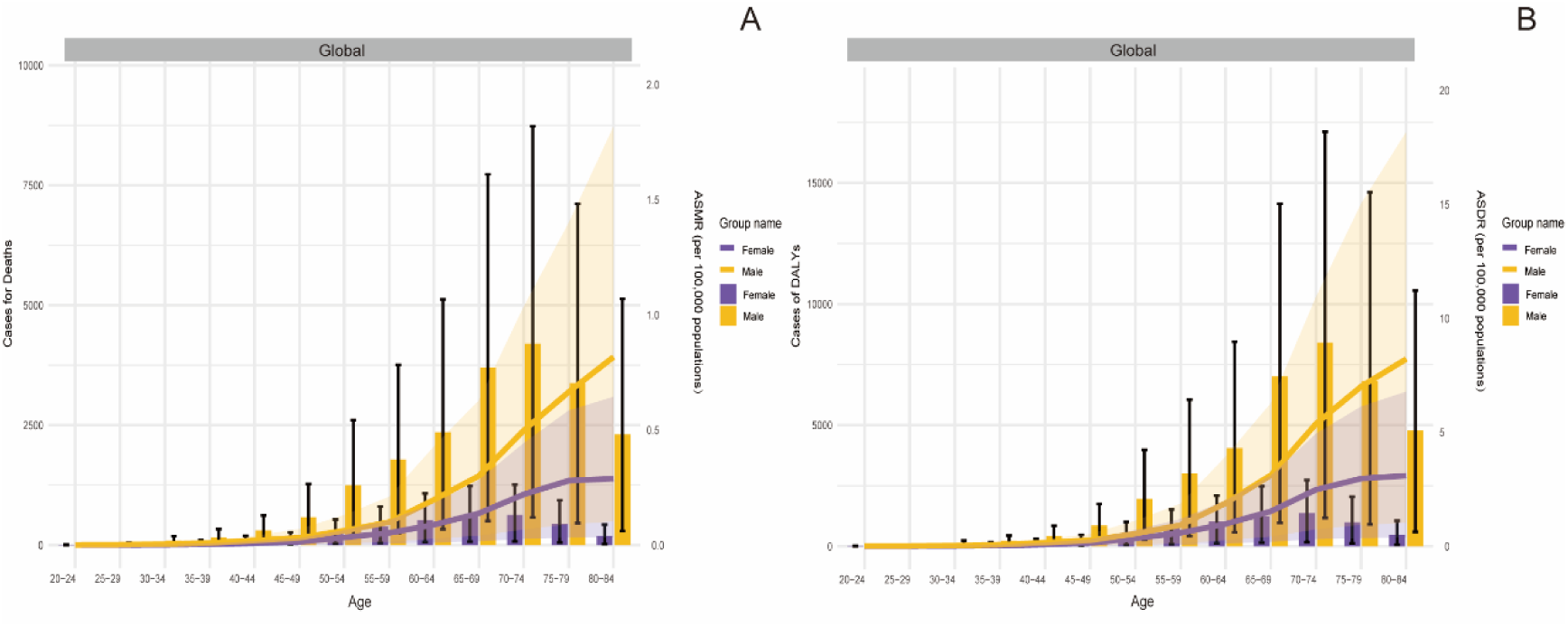
Cases and ASR of IS-HBMI by different sexes in 2021. Notes: (A) deaths, (B) DALYs. IS-HBMI, ischemic stroke attributable to high body-mass index; DALYs, the Disability-Adjusted Life Years; ASR, age-standardized rate; ASDR, age-standardized DALYs rate.

Time trend analysis showed that the average annual percentage change (AAPC) in ASR between 1990-2021 was −0.92 (95% CI: [−1.07 to −0.77]) for females, which was significantly better than that of males(−0.1, 95% CI: −0.21 to 0, p<0.001) for difference between genders) (fig 2). There was significant regional heterogeneity in this gender difference, which was particularly prominent in high SDI areas showed a significant decline in ASR AAPC (−1.49), with females maintaining a better improvement trend (−0.92) consistent with the global pattern, suggesting that the female group had greater risk improvement in medically well-resourced areas. These results suggest that there is a gender asymmetry in the effectiveness of IS-HBMI prevention and control, and that it is closely associated with the level of socioeconomic development.

**Fig 2.**
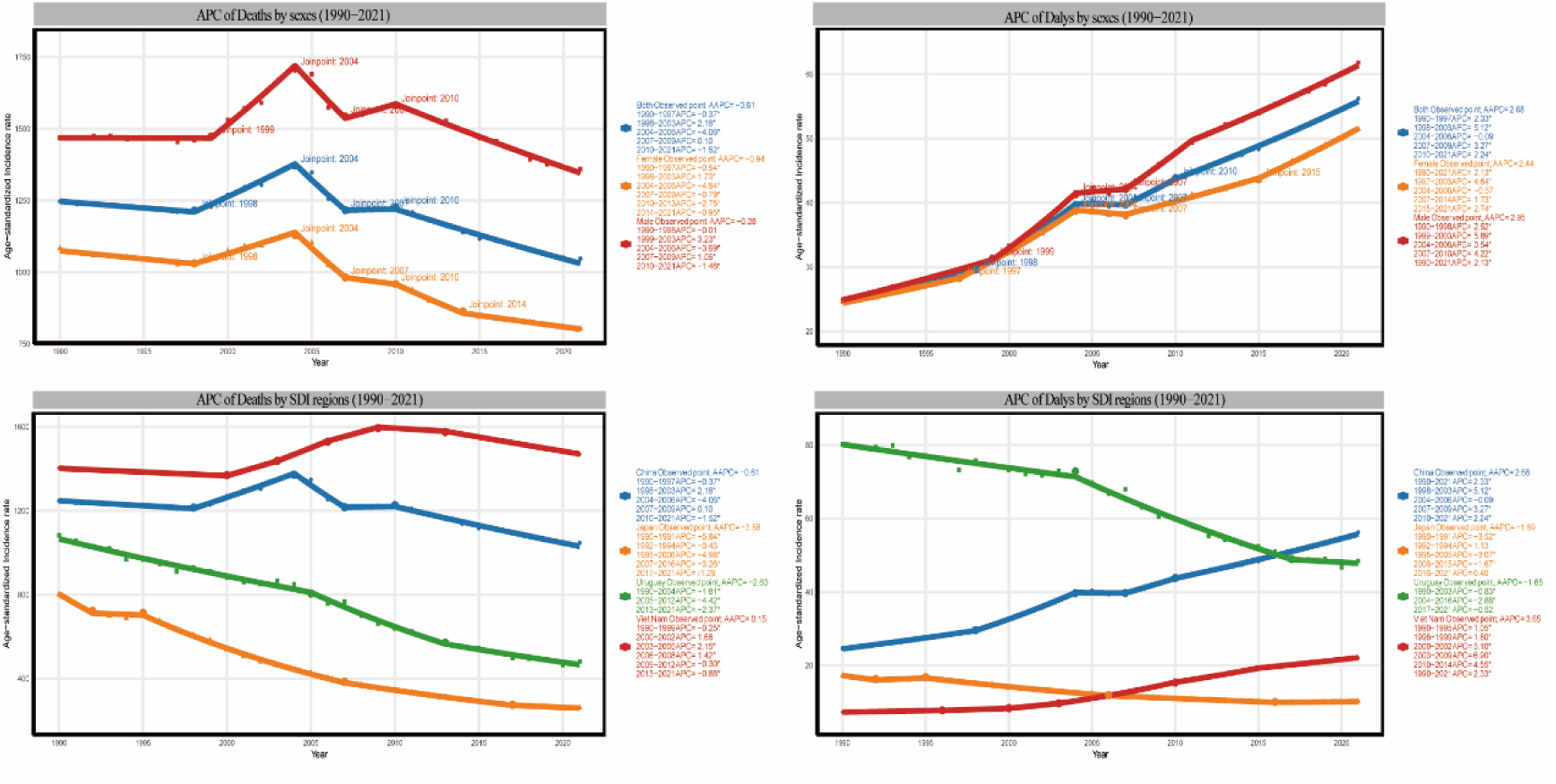
Trends of IS-HBMI from 1990 to 2021, calculated by Joinpoint regression. (A, B) Trends by different sexes for deaths and DALYs, respectively. (C, D) Trends by SDI regions for deaths and DALYs, respectively. IS-HBMI, ischemic stroke attributable to high body-mass index; DALYs, disability-adjusted life years; AAPC, average annual percent change.

### 2. Disease burden of IS-HBMI by different age groups in 1990 to 2021

The global burden of disease for IS-HBMI in 2021 showed significant age stratification, and although the absolute number of deaths was concentrated in the older age groups(fig.6), with the 70-74year olds (711,000), 65-69year olds (678,000), and 75-79year olds (685,000) together accounting for 65.8% of the total number of deaths and constituting the core burden group, the pattern of the age burden varied across regions(fig 6).

The age-standardized mortality rate (ASMR) increased exponentially with age, from 1.21 per 100,000 population in the 20 – 24-year-old group to 287.45 per 100,000 population in the 95 and over group (237.5-fold increase), and regional differences widened with age, with the ratio of the ASMR in high SDI to low SDI areas decreasing from 0.87 in the 20 - 24year old group to 0.23 (p<0.001).

The analysis of time trends showed a pattern of ‘fast decline in old age and slow decline in young age’. Time trend analysis showed that the AAPC showed a pattern of ‘fast decline in the old age and slow decline in the young age’: the young age group (20-29 years old) had a slow decline (AAPC=-0.32 to −0.41), the old age group (70-79 years old) had a significant decline (AAPC=-2.15 to −2.08), and the group of 60-69 years old had a stable trend (AAPC=-0.05 to 0), suggesting that the intervention was effective in the older age group, but that metabolic risk control in the younger age group needs to be strengthened, and that regional inequalities increase with age.

### 3. Disease burden of IS-HBMI by different regions in 1990 to 2021

The global IS-HBMI disease burden in 2021 showed significant socioeconomic development gradient differences. Stratified analyses by SDI showed that although differences in age standardised mortality rates (ASMR) by stratum were relatively small (range 16.36-45.39/100,000 population), the temporal trend diverged extremely significantly: the SDI-stratified AAPC of IS-HBMI ASDR showed significant differences: high SDI (−1.49), high-middle SDI (−1.46), middle SDI (1.21), low-middle SDI (1.59), and low SDI (−1.47). Notably, middle SDI and low-middle SDI regions showed an upward trend (AAPC>1.0), the difference being statistically significant (p<0.001).

The GBD regional analyses further reveal extreme divergences: high-income Asia-Pacific (ASDR=11.47/100,000 population, AAPC=-2.14) and Western Europe (consistent with high SDI regional trend, AAPC=-1.49) are characterised by a ‘low-burden, high-decline’ pattern, whereas Southern Sub-Saharan Africa (ASDR=100.52/100,000 population, AAPC=1.93) showed an upward trend, consistent with the middle and low-middle SDI regions(fig 3., Table 2). Among GBD regions, High-income Asia Pacific (represented by Singapore) achieved the lowest ASDR (11.47/100,000) and a significant decline (AAPC=-2.14); Southern Sub-Saharan Africa (represented by Lesotho) showed a high burden (ASDR=100.52/100,000) with an upward trend (AAPC=1.93)(fig 5., Table 2).

**Fig. 3.**
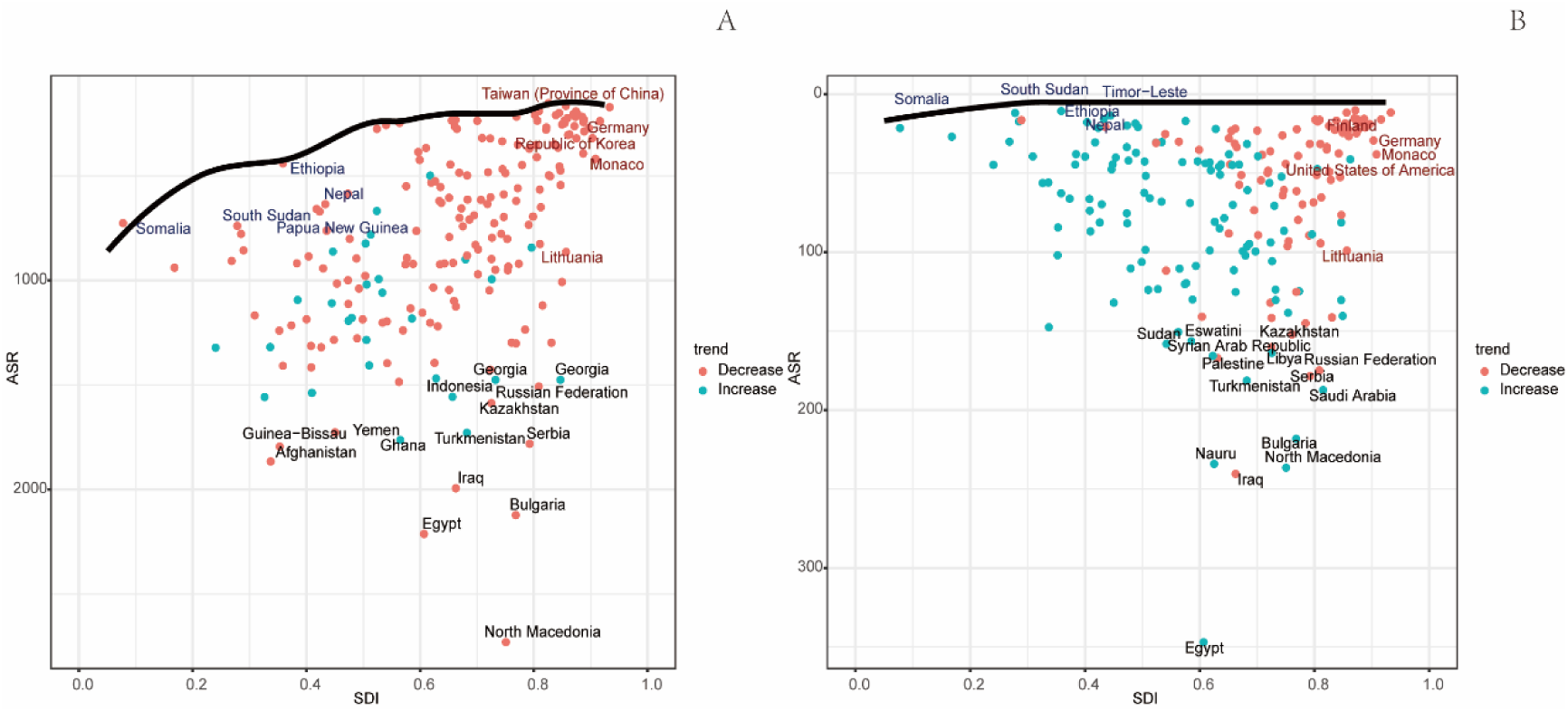
Frontier Analysis of Age-Standardized Mortality Rates for Ischemic Stroke in 2021 (Classified by Multiple Risk Factors and High Body Mass Index (HBMI)) Each data point represents a specific country or region, while the frontier line indicates the lowest achievable age-standardized mortality rate at a given socioeconomic development level. Countries/regions are plotted based on their Socioeconomic Development Index (SDI) and age-standardized mortality rate. Countries with lower Socio-Demographic Index (SDI) and minimal deviation from the frontier are highlighted in blue, while those with higher SDI and significant deviation relative to their development level are highlighted in red. Data point colors indicate trends in age-standardized rates (ASR) from 1990 to 2021. DALYs refer to disability-adjusted life years; HBMI denotes high body mass index.

**Fig. 4.**
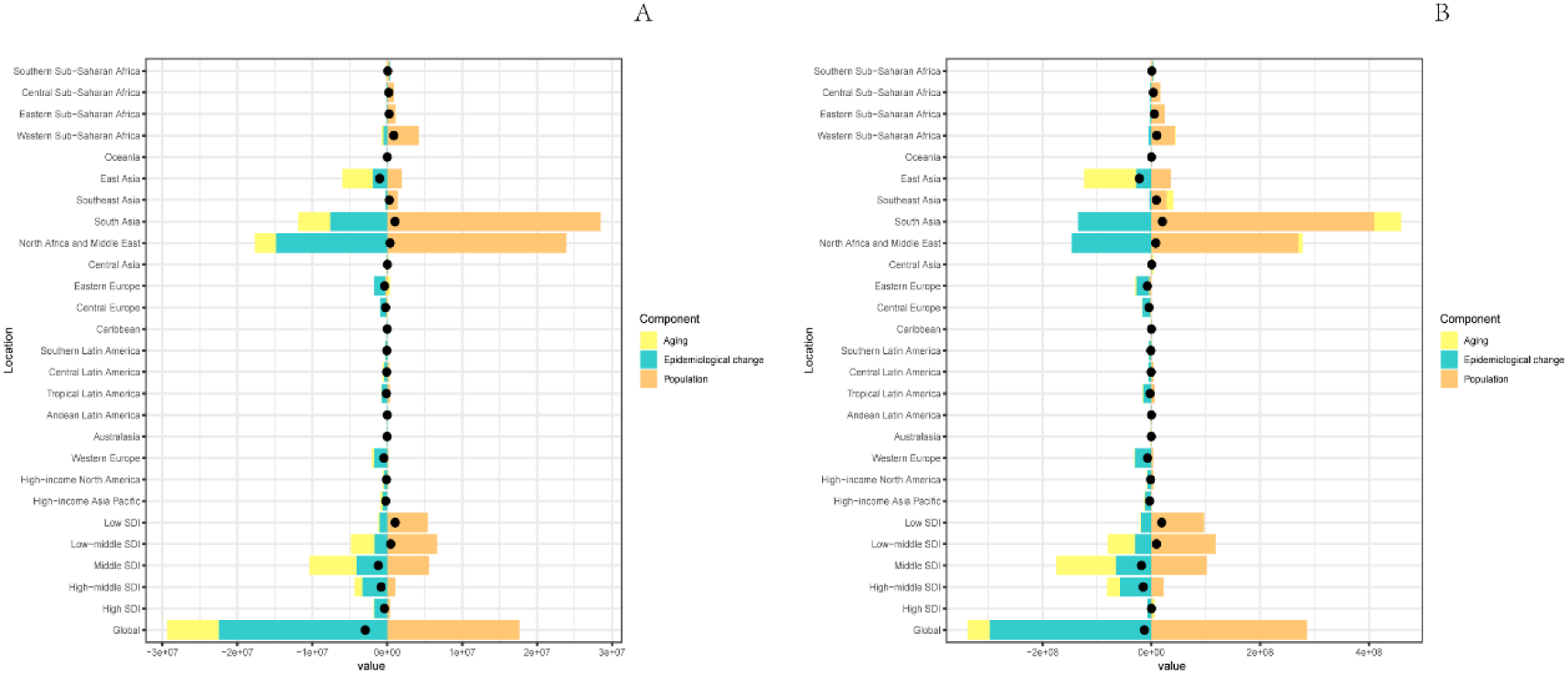
Decomposition analysis in deaths (A) and DALYs (B) of IS-HBMI from 1990 to 2021 by SDI regions and GBD regions. IS-HBMI, ischemic stroke attributable to high body-mass index; DALYs, the Disability-Adjusted Life Years; SDI, socio demographic index.

**Fig 5.**
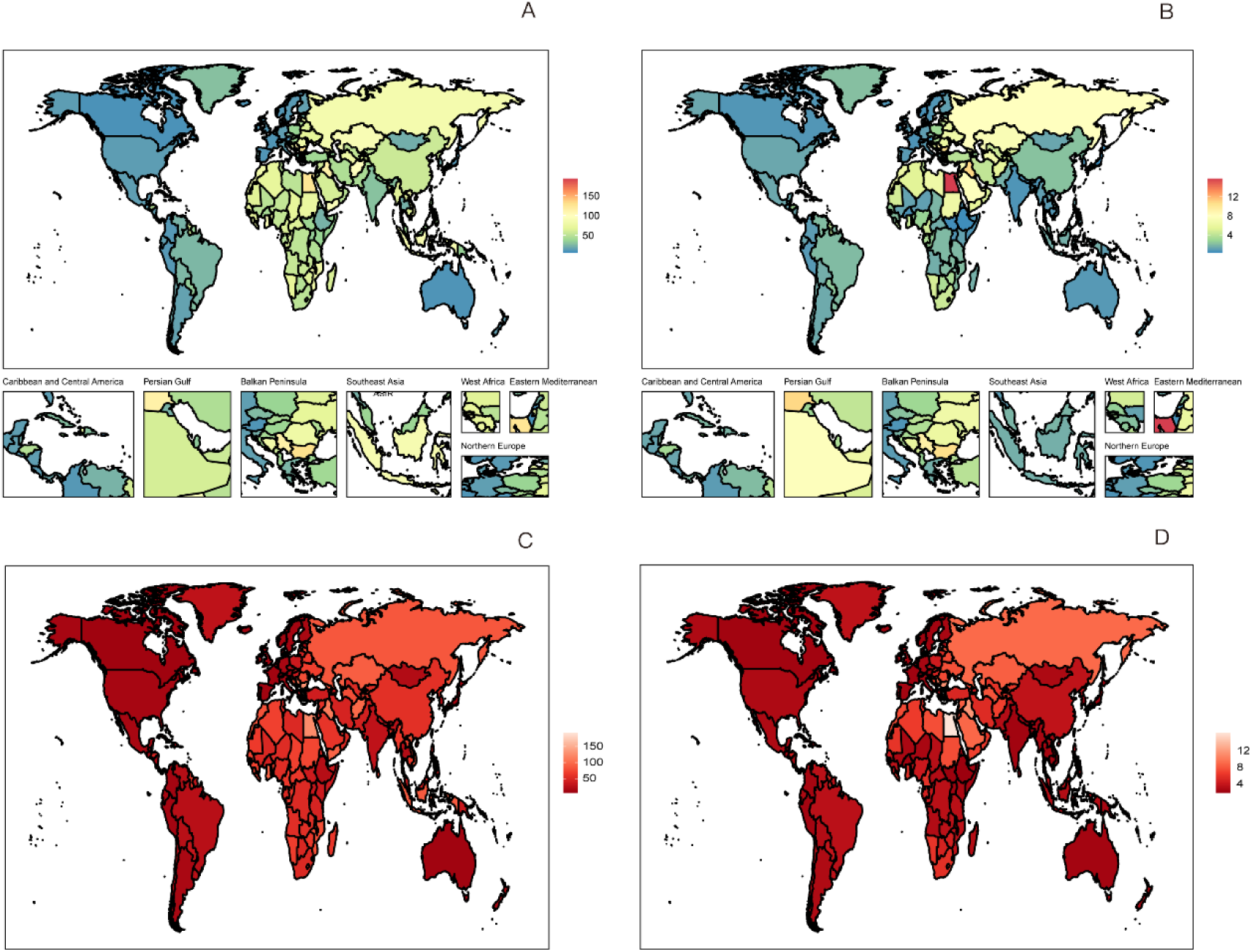
National Burden of Ischemic Stroke Attributable to High Body Mass Index, 2021 Age-standardized rates (A, B) and their average annual percentage change (C, D). Abbreviations: AAPC, average annual percentage change; ASDR, age-standardized DALY rate; ASMR, age-standardized mortality rate; ASR, age-standardized rate; DALYs, disability-adjusted life years; HBMI, high body mass index; IS, ischemic stroke.

China, as a representative of the medium-high SDI region, has achieved remarkable results in controlling the burden of IS-HBMI. Through the implementation of special initiatives such as ‘Healthy China 2030’, China has achieved a rapid decline in ASMR (e.g., APC = −2.00 for the period 2017-2021).Compared to the global trend, China’s rate of improvement is much faster than the global average (AAPC = −0.58) and more significantly better than other medium and high-SDI regions(AAPC=-1.46), which has allowed it to gradually move from being a high-burden country to a low-burden level of a high-SDI country. The case of China proves that strong policy drivers and multisectoral collaboration can effectively curb the rapid growth of IS-HBMI burden. Low SDI blue data points (e.g., Somalia) approach optimal prevention levels with limited resources, while high SDI red data points (e.g., some European countries) deviate from the frontier, indicating room for improvement(fig 3).

### 4. The Relative Importance of High Body Mass Index: A Comparative Analysis Based on the Global Burden of Disease

By comparing high body mass index (HBMI) with the total burden of ischemic stroke attributable to all risk factors, this study reveals the increasing relative importance of HBMI. In 2021, approximately 6.02% of ischemic stroke deaths and 8.28% of DALYs worldwide were attributable to HBMI as a single risk factor, indicating its significant share (table 1, table 2). More critically, the age-standardized DALY rate (ASDR) attributable to HBMI declined at a much slower pace (AAPC = −0.58) than the combined rate for all risk factors (AAPC = −1.62) between 1990 and 2021. This indicates that HBMI’s relative weight within the ischemic stroke risk profile is steadily increasing. This “trend difference” exhibits marked spatial heterogeneity, most pronounced in medium-SDI regions undergoing rapid socioeconomic transformation. In these areas, the ASDR for IS-HBMI even shows an upward trend (AAPC = 1.21), starkly contrasting with the overall improvement in disease burden. In summary, comparative analysis confirms that despite global progress in controlling other risk factors, obesity prevention and control efforts have lagged behind(fig 6). Consequently, the HBMI has emerged as an increasingly influential driver, underscoring the urgent need for targeted prevention and control strategies in the future.

**Fig 6.**
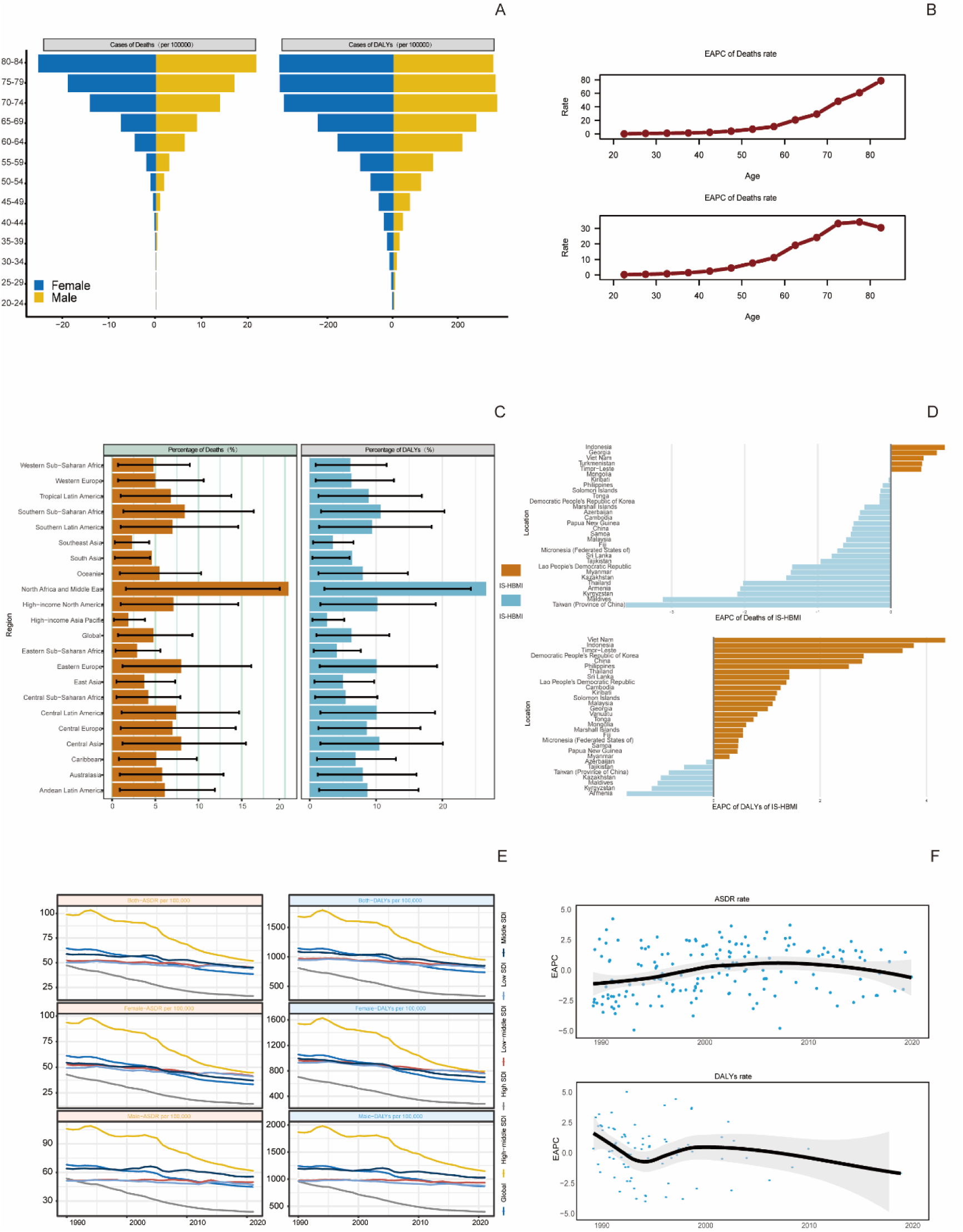
Burden of ischemic stroke attributable to high body-mass index (IS-HBMI): deaths, disability-adjusted life years (DALYs), and their average annual percentage change (AAPC), 1990–2021. Results are shown by age (A, for burden; B, for AAPC), by Socio-demographic Index (SDI) quintile and Global Burden of Disease (GBD) region (C), and by sex over time (E).

**Table 1.**
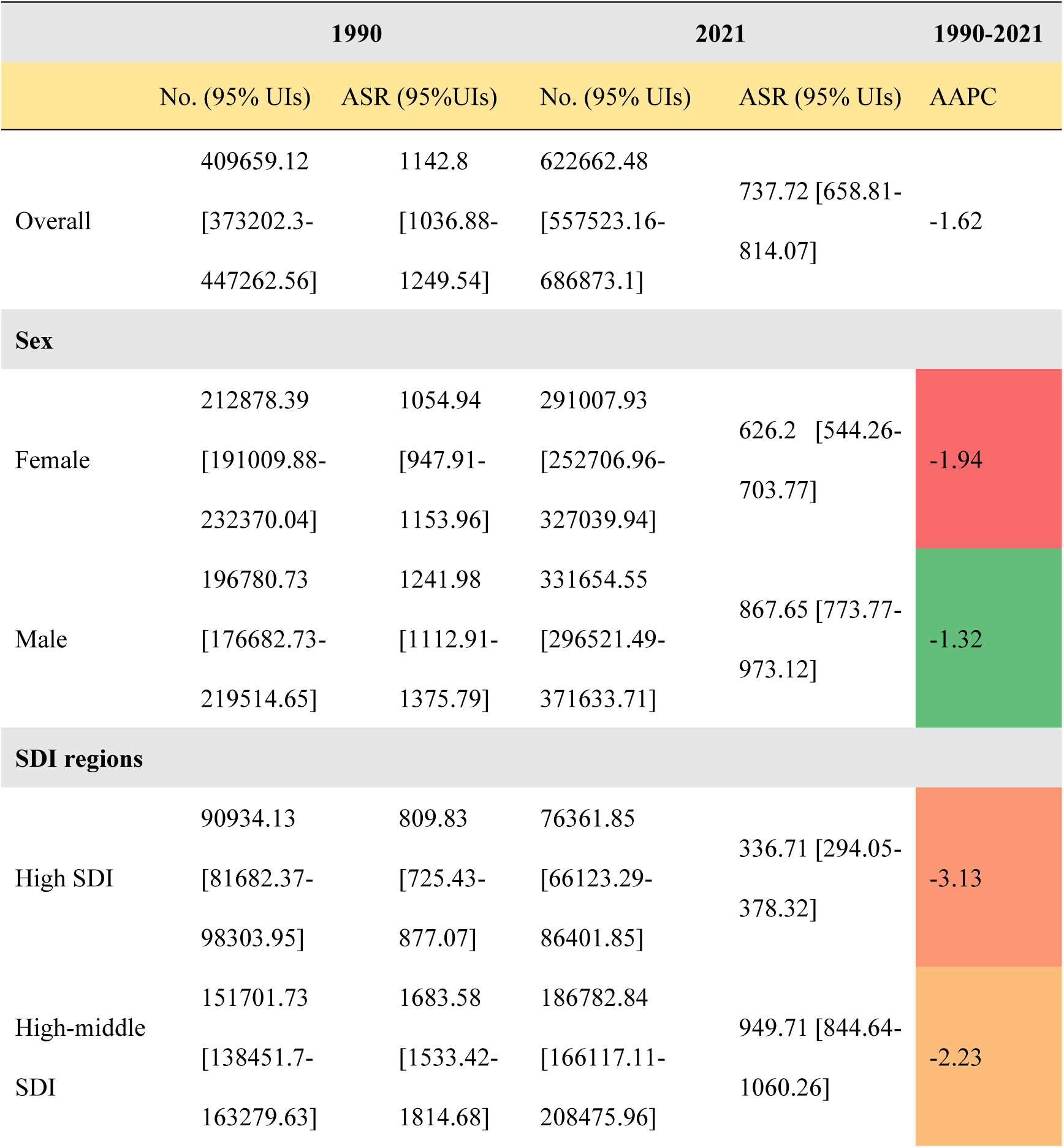
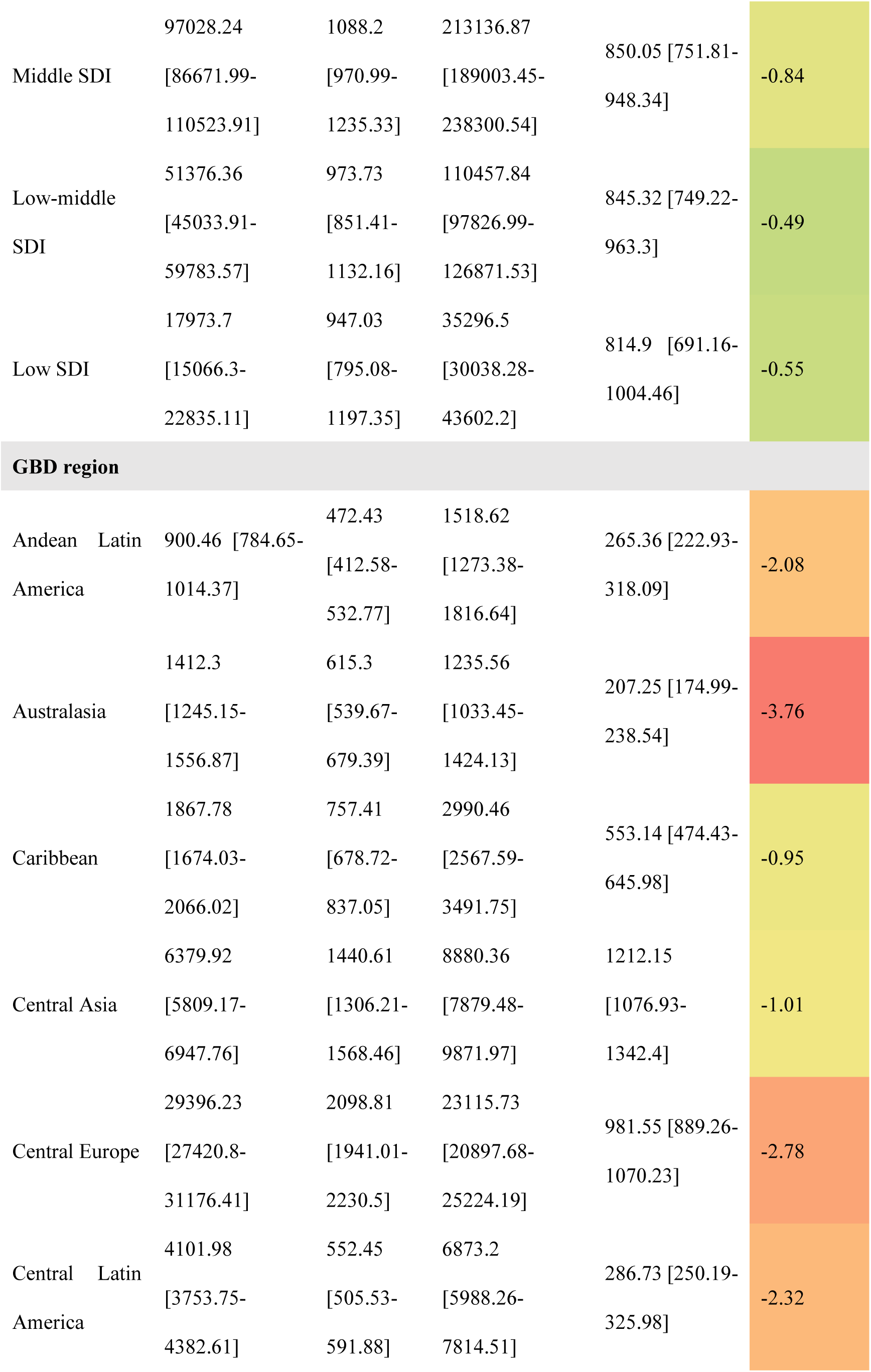
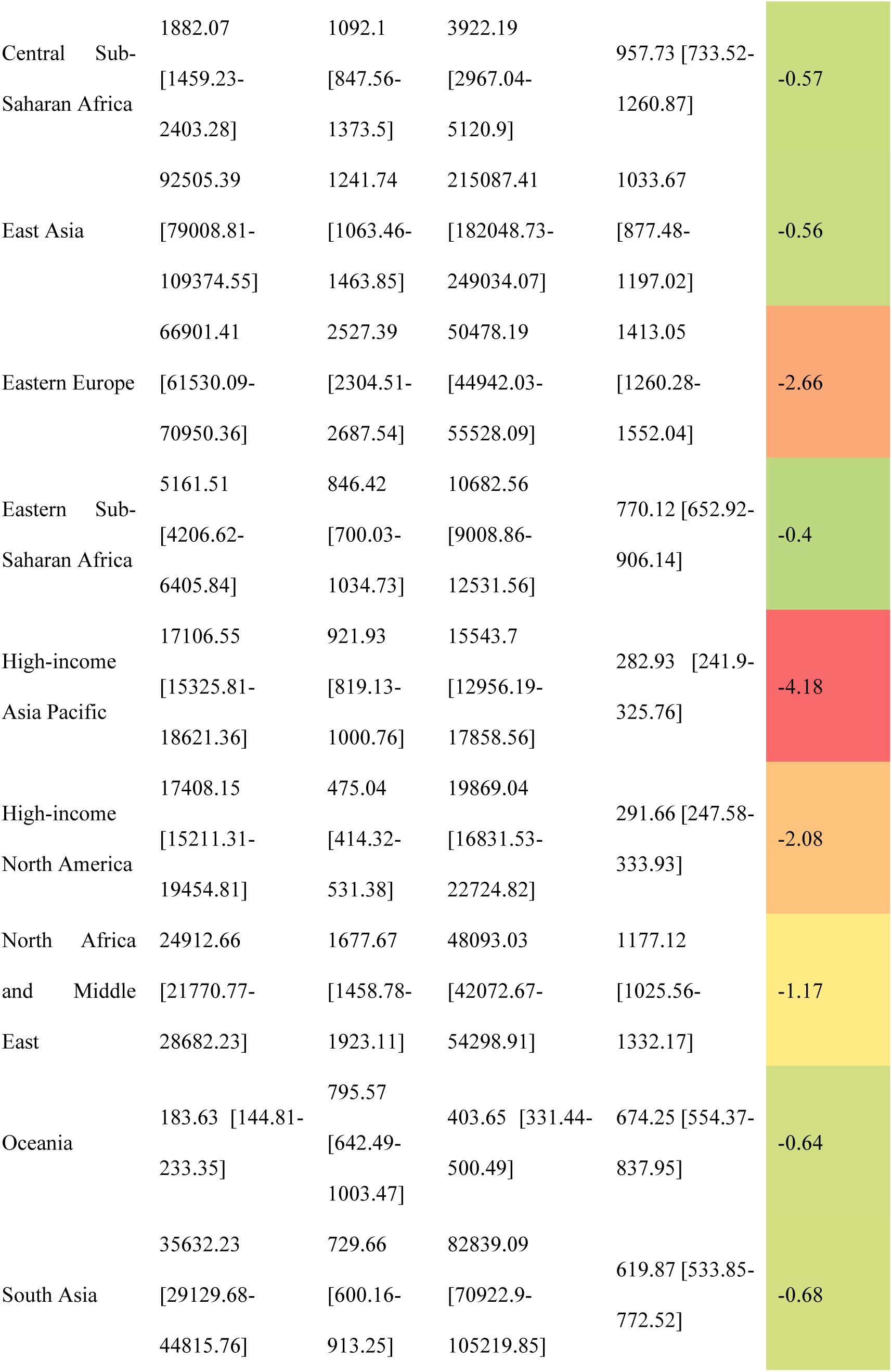
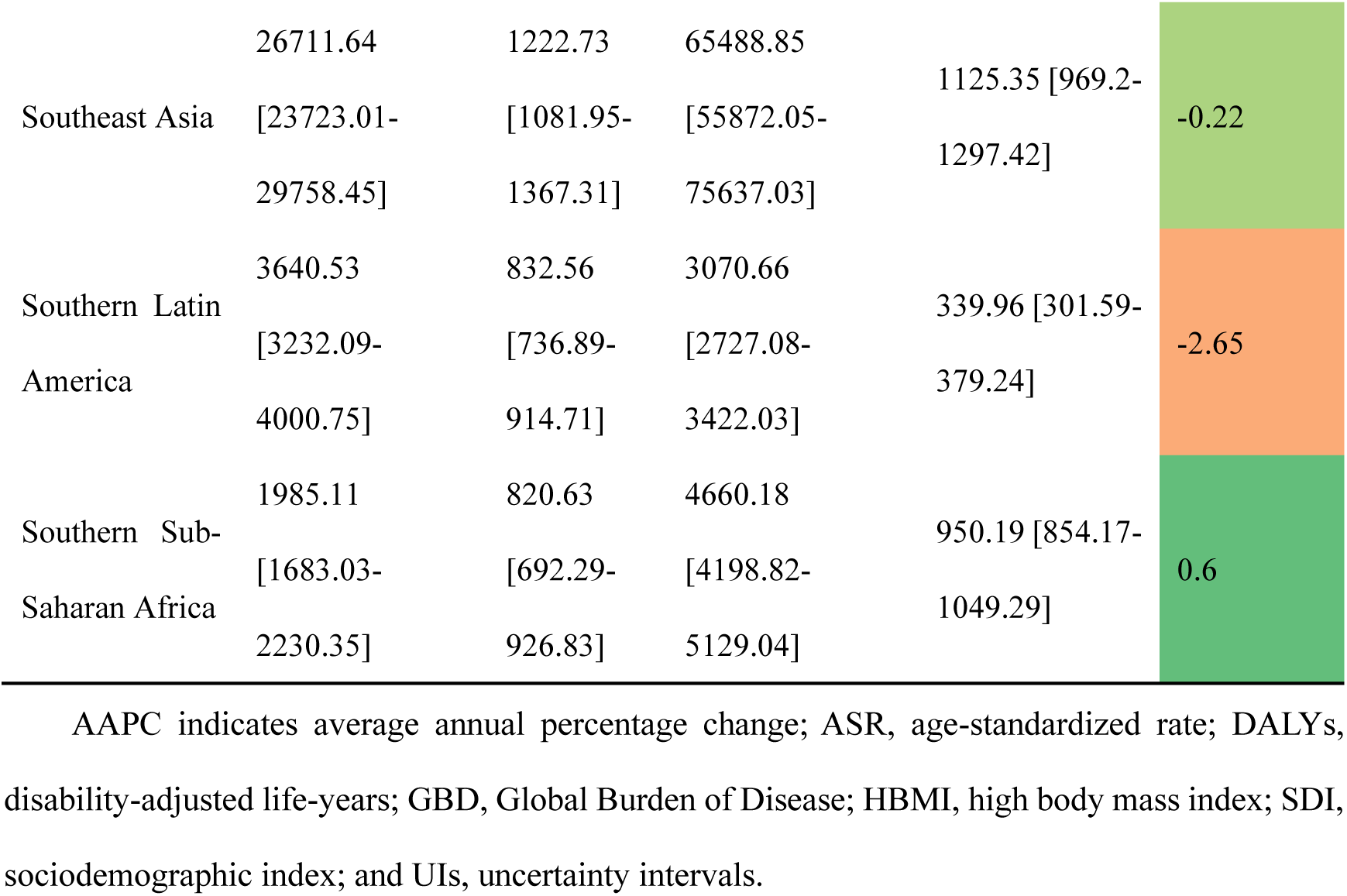
Number of Cases and Age-Standardized Rate (ASR) of Disability-Adjusted Life Years (DALYs) Attributable to Multiple Risk Factors for Ischemic Stroke, by SDI Quintile and GBD Region, in 1990 and 2021 (with Annual Percentage Change), and the Average Annual Percentage Change (AAPC) from 1990 to 2021.

**Table 2.**
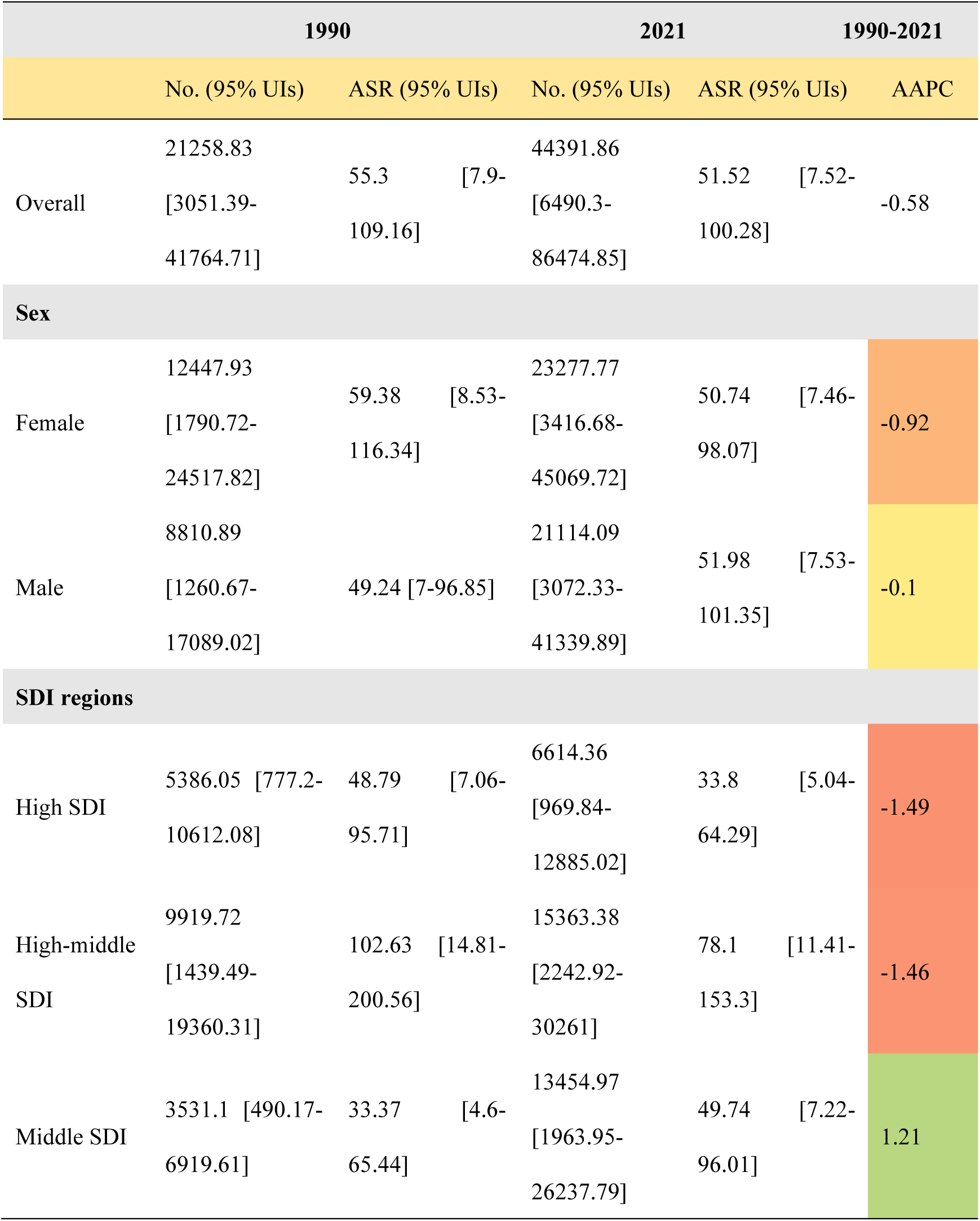
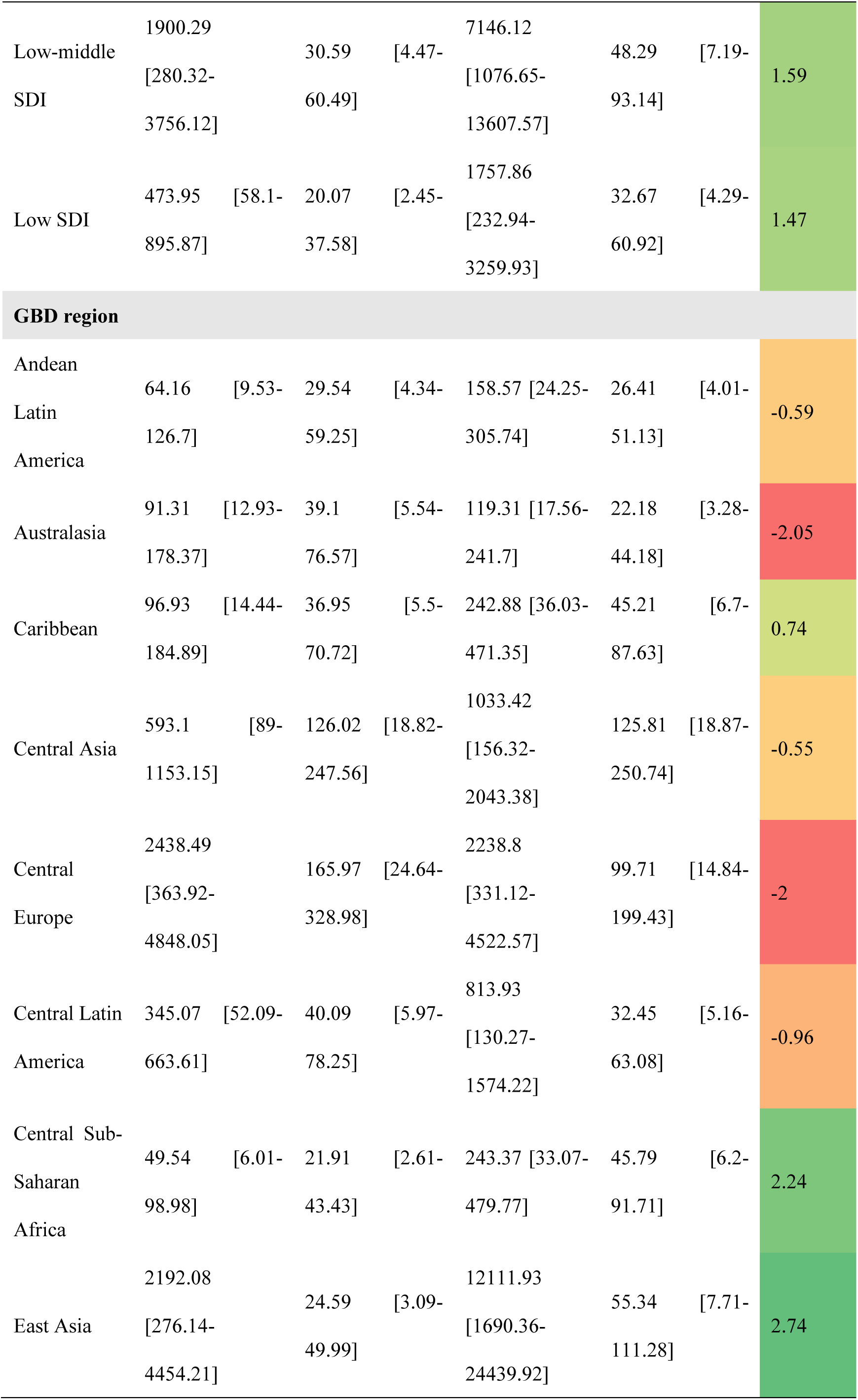
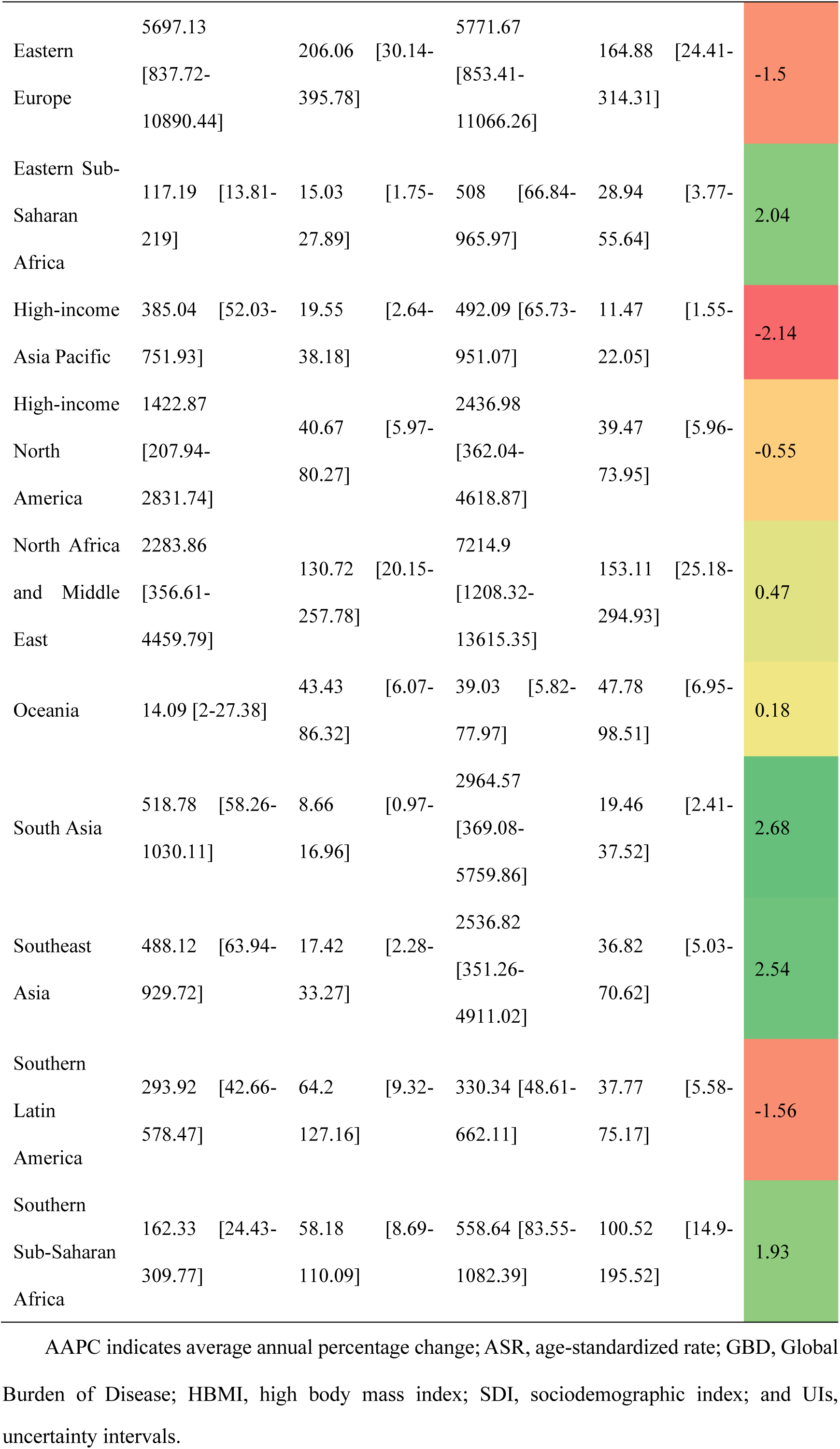
Case Number and ASR of DALYs of ischemic stroke Attributed to HBMI in 1990 and 2021 by different sexes, SDI Quintiles and GBD Regions, With AAPC From 1990 to 2021. Note: All numerical values in this table are scaled down by a factor of 100 (i.e., original values divided by 100) for presentation clarity.

### 5. Projection of IS-HBMI disease burden from 2022 to 2050

Projections for 2022 to 2050 suggest that the global IS-HBMI age-standardized mortality rate (ASMR) will end a period of rapid decline and enter a plateau of near-zero increases, but this general plateau hides growing health inequalities between regions(fig 7.).

**Fig 7.**
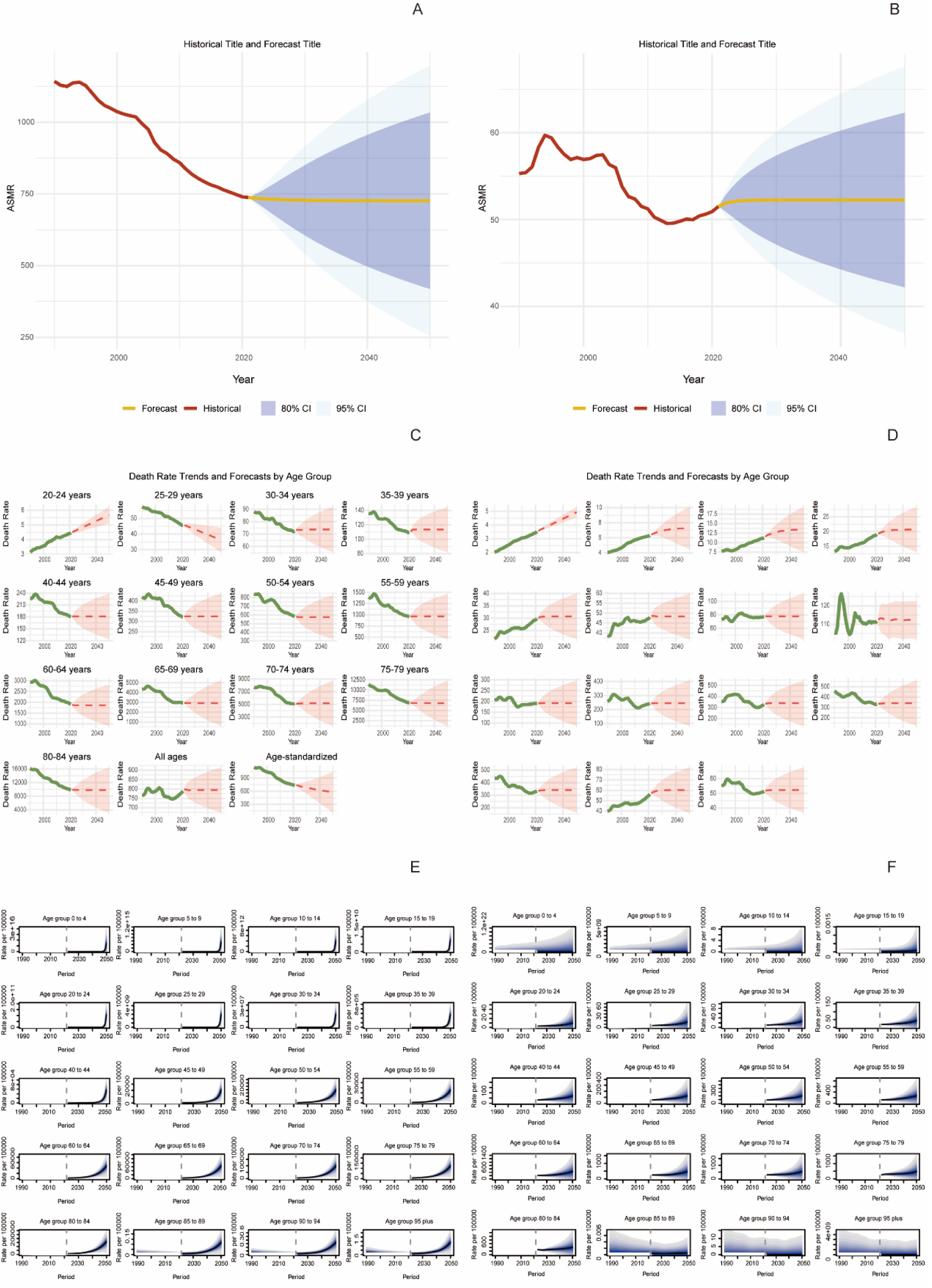
Historical Projected Number of Cases and Age-Standardized Rates of DALYs and Mortality for Ischemic Stroke Attributable to Multiple Risk Factors(A, C, E) and to High Body Mass Index(B, D, F) through 2050 Abbreviations: ASDR, age-standardized disability-adjusted life years rate; ASMR, age-standardized mortality rate; DALYs, disability-adjusted life years; HBMI, high body mass index; IS, ischemic stroke.

Specifically, ASMR in high socio-demographic index (SDI) regions (e.g., Western Europe) is projected to decline steadily from 13.97/100,000 population to 11.5/100,000 population, demonstrating its strong disease control, while China, as a model for medium-high SDI regions, is projected to experience a significant decline in ASMR from approximately 38/100,000 to 25.1/100,000 through active interventions, with the rate of decline (34%) far exceeding the global average. However, the outlook for low-SDI regions is extremely grim, with the ASMR projected to have risen from 52.8 to 58.1 per 100,000, resulting in a dramatic widening of the mortality gap between high- and low-SDI regions from 27.55 to 46.6 per 100,000, and a deepening of the inequality gap by nearly 70 per cent.

At the same time, age-stratified projections reveal inherent risks: the middle-aged group (45-64 years) will see a steady decline in burden (e.g., from 35/100,000 to 20/100,000 in the 55-59 year old group), which is a reliable focus for prevention and control; the young group (20-44 years) has low mortality, but very wide confidence intervals (e.g., 95% UIs: 0.2-0.6/100,000 in the 30-34 year old group) suggesting great uncertainty associated with the younger age group for obesity; and the elderly group (≥65 years) will see limited improvement (e.g., less than 25% in the 80-84 year old group), whose high and persistent burden will continue to challenge healthcare systems. and limited improvement (e.g., less than 25% reduction in the 80-84 age group) in the older age group (≥65 years), whose high and persistent burden will continue to challenge the healthcare system.

In summary, the future global burden of IS-HBMI is shifting from a generalised improvement to an intensification of differentiation, with low SDI regions emerging as new growth poles and the accumulation of risk early in the lifecycle posing a long-term threat that urgently requires more targeted and equitable global prevention and control strategies. A more targeted and equitable global prevention and control strategy is urgently needed.

## Discussion

This study provides a comprehensive analysis and projection of the global burden of disease for IS-HBMI.The global burden of IS-HBMI has continued to increase from 1990 to 2021. Much of this increase is attributed to breakthroughs in diagnostic tools and increased awareness of screening[27]. A systematic analysis of global burden of disease data from 1990 to 2021 reveals the stark current state of the burden of ischemic stroke due to high body mass index, significant disparities and future challenges[28, 29]. Our analysis confirms that, despite global progress in age-standardized mortality, the absolute number of deaths from IS-HBMI continues to rise, constituting a growing global public health burden. This divergence between ‘rising absolute numbers and falling standardised rates’ reflects the complex interplay of population growth, ageing and epidemiological shifts.

One of the most striking findings of this study is the existence of large inequalities in IS-HBMI burden, which are manifested in three main dimensions: socioeconomic, gender, and age. First, socioeconomic inequality was the central driver. The analysis reveals a convergence in the reduction of disease burdens between high-SDI (AAPC: −1.49) and low-SDI (AAPC: −1.47) regions. In contrast, the upward trends in middle-SDI (1.21) and lower-middle-SDI (1.59) regions signify a concerning divergence. This points to a reconfiguration of global health inequalities, where the traditional wealth-health gradient is being complicated by the relative decline of middle-income countries, even as the lowest-income countries show progress. High SDI regions, with their well-established healthcare systems, advanced screening technologies and high public health literacy, have succeeded in significantly reducing the risk of age-standardized[30]. In contrast, although low-SDI regions overall showed a mild decline (AAPC=-1.47), subregions such as Southern Sub-Saharan Africa are facing a ‘double burden of disease’ of malnutrition and obesity epidemics, with their health systems weakly equipped to cope, leading to a rising burden of IS-HBMI (AAPC=1.93)[31, 32]. This SDI gradient is even more pronounced at the regional and national levels, ranging from low burden, high rate of decline in the high-income Asia-Pacific region to high burden, positive rate of growth in Lesotho, a stark contrast that underscores the deterministic nature of the burden of disease at the level of economic development Impact[33, 34]. Secondly, analysis revealed significant gender differences with distinct regional patterns. Although the age-standardized rate (ASR) of IS-HBMI was higher in men (51.98 per 100,000) than in women (50.74 per 100,000), the trend showed a stark reversal: the average annual percentage change (AAPC) indicated a considerably better rate of improvement for women (−0.92%) compared to men (−0.1%)[35, 36]. This disparity in progress, most evident in high-SDI settings, implies that women might achieve better outcomes when healthcare resources are secured. Consequently, intervention strategies should be sex-specific, addressing risk factors predominantly in men while leveraging protective factors in women[37]. Thirdly, age-stratified analyses reveal the transfer of risk over the life cycle: while the older age group remains at the centre of the absolute burden, the trend of a slow decline in ASMR in the younger age group (20-29 years) (AAPC=-0.32∼-0.41) is particularly worrying[38]..This implies that the long-term metabolic risks associated with exposure to high BMI in early life stages are accumulating and may set the stage for future disease outbreaks.[39, 40]. The significant improvement in the burden in the older age group is evidence of the effectiveness of current interventions for this population. At the same time, the significant improvement in the burden in the older age group is evidence of the effectiveness of current interventions targeting this population, but its high and persistent absolute burden will continue to challenge the health-care system[41, 42].

The analysis of the Chinese case provides valuable lessons for intermediate SDI countries. Through the implementation of strong national policies, such as Healthy China 2030, China has achieved a rapid decline in ASMR, proving that policy-driven and multisectoral collaboration in the process of economic development can effectively curb the rapid increase in the burden of IS-HBMI, which provides a model to be followed by countries facing similar challenges[43].

The results of forecast modelling for the future to 2050 are a wake-up call. Global ASMR enters a plateau, masking growing regional divergence[44]. Low-SDI regions are projected to be the new pole of burden growth, while the burden in high-SDI regions will continue to decline, leading to a further deepening of the global health divide by nearly 70 per cent. In addition, the wide confidence intervals of the projections for the youth group hint at the great uncertainty associated with the rejuvenation of obesity, posing a potential threat in the long term[45]. In summary, the findings of this study highlight the need to move from a ‘one-size-fits-all’ universal strategy to a precise and stratified global prevention and control.

Taken together, the findings of this study highlight the need to shift from a one-size-fits-all universal strategy to a precise, stratified global prevention and control approach to address the challenge of IS-HBMI. For High SDI regions, the focus should be on sustaining existing gains and intervening in adolescent obesity; for medium and High SDI regions, policy-driven multisectoral collaboration needs to be strengthened to control metabolic risks associated with economic development; and for low SDI regions, the international community should prioritise assistance in establishing an integrated prevention, screening, and treatment system with low cost. For low SDI regions, the international community should give priority to assisting them in establishing an integrated ‘prevention-screening-treatment’ and low-cost early prevention and control system, so as to prevent IS-HBMI from becoming a new challenge that is beyond their capacity[46]. Future research should aim to elucidate the long-term effects of high BMI exposure in adolescence and develop optimised prevention and control programmes based on cost-effectiveness to ultimately contribute to an equitable decline in the global burden of IS-HBMI.

### Conclusion

This study shows that the global burden of ischemic stroke due to high body mass index increased significantly in absolute terms between 1990 and 2021, with a 152.6% rise in deaths. However, age-standardised rates (ASR) showed a decreasing trend. In contrast to the sustained burden in low-SDI regions, high- and middle-high-SDI regions achieved pronounced reductions in ASR, with AAPCs of −1.49 and −1.46, respectively. Significant differences were also observed across gender and age dimensions. ASR was consistently higher in men than in women, but improved more rapidly in women, particularly in high-SDI areas. The burden of disease remains concentrated in older age groups, but the rate of decline has slowed in younger age groups, suggesting that the risks associated with early-life obesity are emerging.

Projections to 2050 indicate that global ASMR trends will level off, but the disparities observed in this study are expected to evolve. The overall modest decline in low SDI regions (AAPC = −1.47) masks a rising burden in subregions such as Southern Sub-Saharan Africa (AAPC = 1.93). This divergence suggests that health inequalities will not only persist but become more complex. In the absence of targeted interventions, the burden will continue to climb in these specific geographic hotspots. These findings underscore the urgent need for equitable and locally tailored strategies to mitigate the growing impact of obesity-related health problems.

## ACKNOWLEDGMENTS

This work was supported by Joint Funds for the Innovation of Science and Technology, Fujian Province (grant/award number: 2024Y9123), and National Natural Science Foundation of China (grant/award number: 82301543).

## CONFLICT OF INTEREST STATEMENT

The authors declare no competing interests.

## DATA AVAILABILITY STATEMENT

The datasets generated during and/or analyzed during the current study are available from the corresponding author upon reasonable request.

## Notes

### Competing Interest Statement

The authors have declared no competing interest.

### Funding Statement

This work was supported by National Natural Science Foundation of China: 82301543. and Joint Funds for the Innovation of Science and Technology,Fujian Province: 2024Y9123,

### Author Declarations

This study involved analysis of existing, de-identified, aggregated data from the Global Burden of Disease (GBD) study, which is publicly available. Therefore, it was considered exempt from review by the Fujian medical university Institutional Review Board (IRB) or Ethics Committee.

